# Mitigation and herd immunity strategy for COVID-19 is likely to fail

**DOI:** 10.1101/2020.03.25.20043109

**Authors:** Barbara Adamik, Marek Bawiec, Viktor Bezborodov, Wolfgang Bock, Marcin Bodych, Jan Pablo Burgard, Thomas Götz, Tyll Krueger, Agata Migalska, Barbara Pabjan, Tomasz Ożański, Ewaryst Rafajłowicz, Wojciech Rafajłowicz, Ewa Skubalska-Rafajłowicz, Sara Ryfczyńska, Ewa Szczurek, Piotr Szymański

## Abstract

On the basis of a semi-realistic SIR microsimulation for Germany and Poland, we show that the *R*_0_ parameter interval for which the COVID-19 epidemic stays overcritical but below the capacity limit of the health care system to reach herd immunity is so narrow that a successful implementation of this strategy is likely to fail, which is in contrast to results obtained from classical differential equation models. Our microsimulation is based on official census data and involves household composition and age distribution as the main population structure variables. Outside household contacts are characterised by an out-reproduction number *R*^*^ which is the only free parameter of the model. For a subcritical domain we compute the time till extinction and prevalence as a function of the initial number of infected individuals and *R*^*^. For the Polish city of Wrocław we also discuss the combined impact of testing coverage and contact reduction. For both countries we estimate *R*^*^ for disease progression until 20th of March 2020.

## 1. Introduction

Mitigation of a novel infectious disease with the aim to reach herd immunity is a classical textbook concept in epidemiology and has been successfully applied in the past, foremost in the case of novel influenza strains^1–3^. The idea is simple: in the absence of a vaccination for a novel infectious disease one tries to flatten the incidence curve to such an extent that the daily number of cases that require medical assistance is kept below the capacity of the health care system. The long term goals are to obtain a sufficiently large fraction of the population that has become infected and to reach herd immunity which would lead to a less severe or even subcritical second outbreak wave. On the other hand, an extinction strategy would aim at introducing sufficient contact reductions to keep the epidemic subcritical and not lifting these restrictions until the disease becomes extinct.

COVID-19 is a novel disease which most likely emerged from a zoonotic event in China at the end of 2019^4^. In the meantime, it has affected more than 170 countries and surpassed 400 000 cases worldwide. The disease is highly infectious and spreads through droplets, similar to influenza. The case fatality rate seems to depend heavily on the quality of treatment and is currently (at the time of writing) estimated to be between 1.4 and 5 percent ^5^. Of particular concern is the large number of patients requiring either breathing assistance or treatment in an intensive care unit (ICU)^6^. Different countries have implemented different defense strategies which are constantly being updated and adapted to the actual prevalence. The decisions mainly are based on SIR or SEIR models and focusing at the reproduction number *R*_0_. For example at http://covidsim.eu/ changing the parameters for Germany, the *R*_0_ can vary more or less between 1 and 1.4 to remain overcritical, but not exhausting the health care system. However, these models do not include one of the most important infection paths, which are the households. A comparison of such a SEIR model and our microsimulation can be found in the Appendix C in Figure C.14.

In this article, we study the likelihood of success of such strategies, based on a semi-realistic microsimulation model for the spread of COVID-19 including household structures. Simulations with census based household compositions and age distribution were carried out for Germany and Poland and for two representative major cities, Berlin and Wrocław. Microsimulations are considered an appropriate tool to describe complex structures of infection paths and disease progression and have been performed in the past for influenza^7^ and recently also for Covid-19^8^. Our model summarises the net effect of all secondary infections caused by an infected individual outside its own household into an out-reproduction number *R*^*^ which is the only free parameter in our model. We assume that the interactions within the household are hardly affected by social distancing strategies. Thus, the *R*^*^ parameter best reflects the strength of non-pharmaceutical interventions. The stronger the interventions, the lower the *R*^*^. The mitigation strategy, however, needs to allow *R*^*^ to be high enough to enable the population to reach herd immunity. We show that the margin of *R*^*^ for which successful mitigation into an overcritical but not ICU capacity-threatening epidemic can be achieved is extremely narrow, implying that this strategy is likely to fail. Moreover, we quantify the average extent to which social contacts have to be reduced in the population to achieve reasonable times for disease extinction. We present estimates in the case of subcritical epidemics for time till extinction as a function of *R*^*^ and the initial number of infected individuals. The time till extinction has direct economic implications and depends strongly on *R^*^,* showing the critical importance of introducing strong social distancing measures. For Germany, at least an 80% reduction of social contacts outside households is required for the epidemic to become subcritical, and with this reduction, the time to disease extinction amounts to more than a year. An extinction time shorter than a year is only possible with a reduction of social contacts by more than 95 %. For the city of Wrocław we additionally discuss the combined effect of testing coverage and social contact reduction. High testing rates - defined here as the rate of uncovering mild cases - and household quarantine of positive cases allow for less stringent contact reduction but are on their own not sufficient to guarantee a subcritical progression of the epidemic. Finally, we estimate *R^*^* for the present situation in both countries under the assumption that no further spread-preventing actions are taken. Comparing the ratio of the present *R^*^* value with the value at which the epidemic becomes subcritical is a good indicator for the strengths of non-pharmaceutical interventions for a successful extinction strategy. For end-prevalence in the overcritical parameter domain we were able to derive theoretical predictions which match very well with the simulation results. The theoretical results show the strong impact which differences in the household size distribution have on the prevalence and demonstrate complementary to the numerical results how sensitive the prevalence depends on *R*^*^ (see Appendix C).

## 2. Model description

We use an individual based SIR model to describe the spread of COVID-19. Compared to classical ODE models of epidemic spread, microsimulations have the advantage of a better representation of epidemiologically relevant heterogeneity in the population. In addition, the description of the individual disease progression can be used to study the impact of complex countermeasures such as extensive backtracking, testing and quarantine. The model is a non-Markov stochastic process in continuous time based on the infection probability of susceptibles in contact with infected individuals. The contact structure outside of households is represented as an inhomogeneous directed random graph where each node corresponds to an infected individual and the degree of that node corresponds to the number of secondary infections created by that individual. The degree itself depends on how long the individual stays infectious (infectivity time).

### Population structure

Our sample population is based on the census data on the census data (2011) as well as more recent official statistics (2019) from Poland^17–19^ and a synthetic reproduction of the microcensus in Germany (2014)^9^ (see Appendix A) and involves age and household composition. For the relative frequencies of the household sizes see Supplement A. Sample populations were used to represent two major cities with typical urban household structures: Wrocław and Berlin. Since we focus on a conceptual question, more detailed structures like spatial assignment, gender, profession or comorbidity relevant health status are omitted.

### Disease progression within patients

The Covid-19 progression within patients is modelled according to the present medical knowledge. The incubation time is assumed to follow a lognormal distribution with median 3.92 and variance 5.516 [lognormal parameters: shape=0.497, loc=0.0, scale=3.923]. The age dependence of the probability to be hospitalised or to have severe progression or to have critical progression with requirement for ICU treatment is given in Table 1.

**Table 1:** Age dependence of the probability to develop a certain level of symptoms. The probability for death was assumed to be 49% within the critical patients.

| Symptoms | Age groups |  |  |  |  |  |
| --- | --- | --- | --- | --- | --- | --- |
|  | 0-40 | 40-50 | 50-60 | 60-70 | 70-80 | 80+ |
| **Asymptomatic** | 0.006 | 0.006 | 0.006 | 0.006 | 0.005 | 0.004 |
| **Mild** | 0.845 | 0.842 | 0.826 | 0.787 | 0.710 | 0.592 |
| **Severe** | 0.144 | 0.144 | 0.141 | 0.134 | 0.121 | 0.101 |
| **Critical** | 0.004 | 0.008 | 0.027 | 0.073 | 0.163 | 0.302 |

The time till hospitalisation from the onset of symptoms is assumed to be Gamma distributed with median 2.31 and variance 8.365 [gamma parameters: shape=1.177, loc=0.0, scale=2.666]^10^ Patients with non severe progression eventually stay at home and the time from onset of symptoms till staying at home is also assumed to be Gamma distributed with median 1.67 and variance 7.424 [gamma parameters: shape=0.874, loc=0.0, scale=2.915]^11^. The maximal duration of the infectious period is assumed to be 14 days^12^.

### Contact structure and infection transport

Within the households we assume a clique contact structure. Empirical studies have shown that a large fraction of secondary infections are taking place within households^13^. We hence assumed that the probability of a household member to become infected by an already infected household member, who is infectious within a time interval of length *T*, scales as 1 − exp(−*T*/*L*), where *L* + 1 is the household size. Here, the time *T* is measured in days. Outside of the households we assume that infected individuals create on average *c · T* secondary infections, given that all contacts of these individuals are susceptible, where *c* is an intrinsic parameter. Note the time *T* being infectious is different for contacts inside and outside the household. The out-reproduction number *R*^*^ is defined as the expectation of *c* · *T*, which is equal to 2.34*c* under our assumptions of disease progression within patients. The actual number of secondary infections of an individual outside the household is assumed to be Poisson distributed with mean (*c* · *T*). The total reproduction number *R*_0_ is given by the sum of *R*^*^ and the number of secondary infections generated inside the household. The duration of the infectivity time T implicitly depends on age. This is due to the fact that infectivity time is reduced for individuals with severe disease progression, as those patients become hospitalized. Severe progression is in turn more probable for older infected individuals. The outside household contact structure was intentionally chosen to be simple in order to have only one relevant and easily interpretable parameter in the model. We do not consider super-spreading events that could enhance the progression of the epidemic. Such events might have a strong impact at the beginning of an epidemic outbreak but, as the number of cases increases, the mean number of secondary infections R will dominate the evolution.

### Testing and quarantine

For Wrocław we included additional model features to study the effect of testing followed by household quarantine in case the testing was positive. We assume that individuals with severe symptoms will always be detected and individuals with mild symptoms will be detected with probability q two days after the onset of symptoms. A detection is followed up by quarantine of the corresponding household with the effect that all out-household contacts by members of those households are stopped. The parameter q can be interpreted as the likelihood that a person with characteristic mild symptoms will be tested for COVID-19. We did not consider the effect of backtracking in this article since it is a prevention strategy mainly applied during the early and final phases of the epidemic.

## 3. Results and discussion

Table 2 shows the intervals of *R_min_ ≤ R*^*^ ≤ *R_max_* which contain the inteval in which a successful overcritical mitigation is possible for both countries and towns. In other words *R_max_* and *R_min_* are upper and lower bounds for a successful mitigation. A detailed description about how to obtain the values in Table 2 can be found in the Appendix A.

**Table 2:** Intervals of *R_min_ ≤ R^*^ ≤ R_max_* for a possible successful overcritical mitigation.

| | Observed data | Intervals of $R^*$ | | Intervals of $R^*$ obtained for the quarantine scenario (detecting 50% of mild cases and 100% of severe/critical) | |
| --- | --- | --- | --- | --- | --- |
| Entity | Predicted $R^*$ | $R_{\min}$ | $R_{\max}$ | $R_{\min}$ | $R_{\max}$ |
| Poland | 3.16 | 0.26 | 0.37 | — | — |
| Germany | 3.04 | 0.37 | 0.42 | — | — |
| Wrocław | — | 0.35 | 0.42<br>+2.9 %/day | 0.84 | 0.96<br>+3.4%/day |
| Berlin | 3.88 | 0.44 | 0.46 | 0.94 | >1.4 |

Here, successful means that even at the peak of the outbreak the epidemic stays below the capacity threshold of intensive care units. Our capacity thresholds for Germany and Poland are based on public statistical sources^14,15^ and on the very moderate assumption that only 80% of the existing ICU places are occupied^16^. The upper bound for *R*^*^ of those intervals is denoted by *R_max_.* This value is transferred into an average per day growth rate of prevalence, as it is reported by most health offices in their daily situation reports. An average per day growth rate was calculated from the first 50 days of the epidemic. We defined *R_max_* as the smallest *R*^*^ value for which 10 sample paths surpassed the ICU threshold within D days. For cities D was chosen to be 200 days and for countries 700 days. The critical value *R_min_* was defined as the largest *R*^*^ < *R_max_* for which the daily incidence at day 200 was below 50% of the initial number *N_o_* of infected (*N_o_* = 100 for Wrocław, *N_o_* = 1000 for Berlin, *N_o_* = 1000 for Poland and *N_o_* = 15000 for Germany). As can be seen from the values in Table 2, all intervals for a successful mitigation are small, which is below 0.11 in units of *R*^*^.

Timelines for the epidemic at the *R_max_* value are given in Figure 1. Differences in the values of *R_max_*, *R_min_* and the actual value of *R^*^* between the two countries and cities are due to differences in the household structures and different capacity thresholds. We also provide the size of the interval for a successful mitigation for a scenario where households with infected members are quarantined with a 50% chance (for details see section model description). This alternative scenario was only studied for Berlin and Wrocław. Although the location of the critical intervals is slightly different in the alternative scenarios, we observe that the size of critical intervals is only marginally affected (see Table 2).

**Figure 1:**
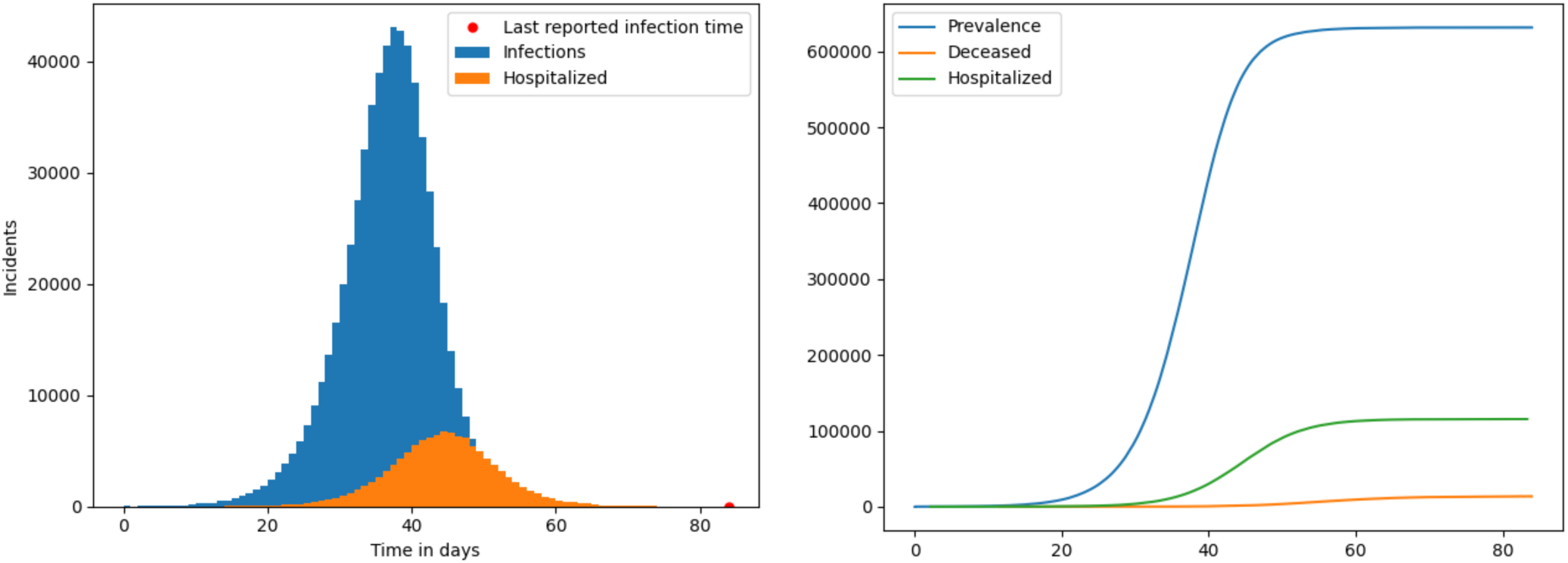
Timeline of the relevant observables for the uncontrolled epidemics: an example outcome of the epidemic in Wrocław growing at *R*^*^ currently observed for Poland starting from 100 infected agents; We run a simulation on a randomly sampled population of 636 thousands of agents that fits the demographics (including. age and household structure) of Wrocław. The left column presents daily incidents: new infections and hospitalization events. The right column shows a plot with the timeline of the epidemic. More than 95% of the population is predicted to be infected within a 3 months time frame starting from the first 100 infected agents.

The present values of *R*^*^ were fitted according to the disease progression till 20th March 2020 in Germany and Poland and summarized in Table 2. In Figure 1 we show the timeline of the relevant observables for the uncontrolled epidemics. For selected values of *R*^*^ in the overcritical domain the end-prevalence of the epidemics for Wrocław are displayed in Figure 2 and Figure 3. We compare them with theoretical predictions from random graph and branching process theory (see mathematical supplement in Appendix C for further details). For values of *R*^*^ within the critical interval Figure 3 shows the level of herd immunity achievable by a successful mitigation.

**Figure 2:**
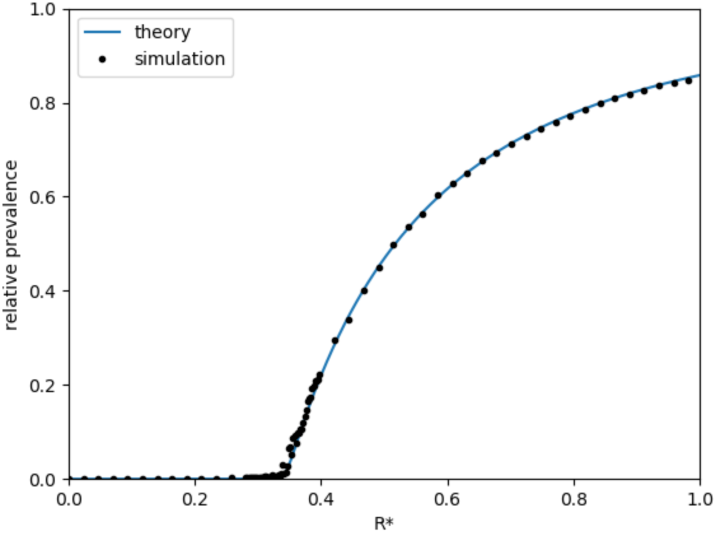
Prevalence as a fraction of whole population in dependence of *R*^*^ for Wrocław

**Figure 3:**
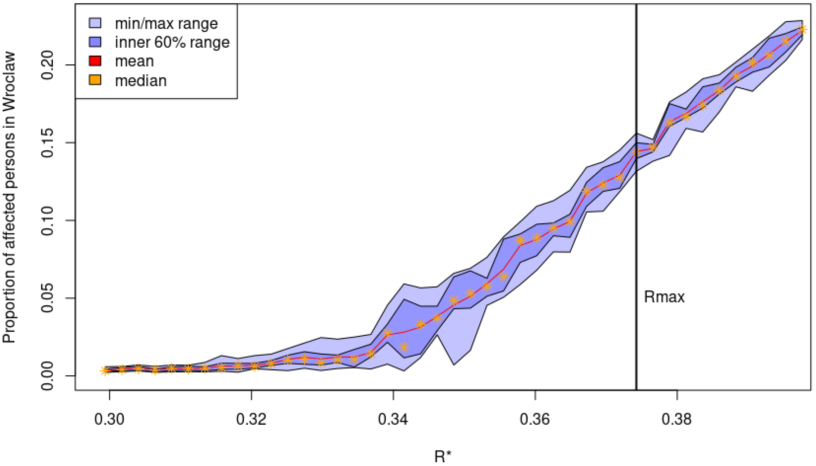
Zoomed region near *R_min_* and *R_max_*.

A heat map for the time till extinction and prevalence in the subcritical domain is given in Figure 4 (detailed numerical values are given in Appendix B). The time till extinction was defined as the first day when no active cases were present. This time is about 1–2 months longer than the time when no new infections appear due to cases with a very long lasting recovery. The extinction times vary strongly in *R*^*^ but only weakly in the initial number of cases. In addition from Figure 4b the dependence of the final prevalence on those two parameters can be obtained.

As can be seen from Figure 4a, extinction times below 8-9 months only occur for values of *R*^*^ below 0.2. In order to put the small values of *R^*^* in the right context and to understand the social implications, one has to compare them to the actual *R^*^* value from Table 2. Till the 20th of March the German growth rate corresponds to the value *R*^*^ = 3.04. A reduction to values less than or equal to 0.2 implicates a 15 fold reduction of social contacts. In other words, out of 15 contacts only one is allowed to persist on average. Hence a lockdown which targets at reasonable low *R*^*^ values has to reduce daily life social contacts (including workplace) by more than 90 %.

Furthermore, we observe that doubling the initial number of infected individuals essentially leads to a doubling of the final prevalence and hence fatalities, independently of the *R*^*^ value. Reported doubling times in many countries are at present between 4-6 days. Therefore an implementation of effective countermeasures as early as possible is recommended.

**Figure 4:**
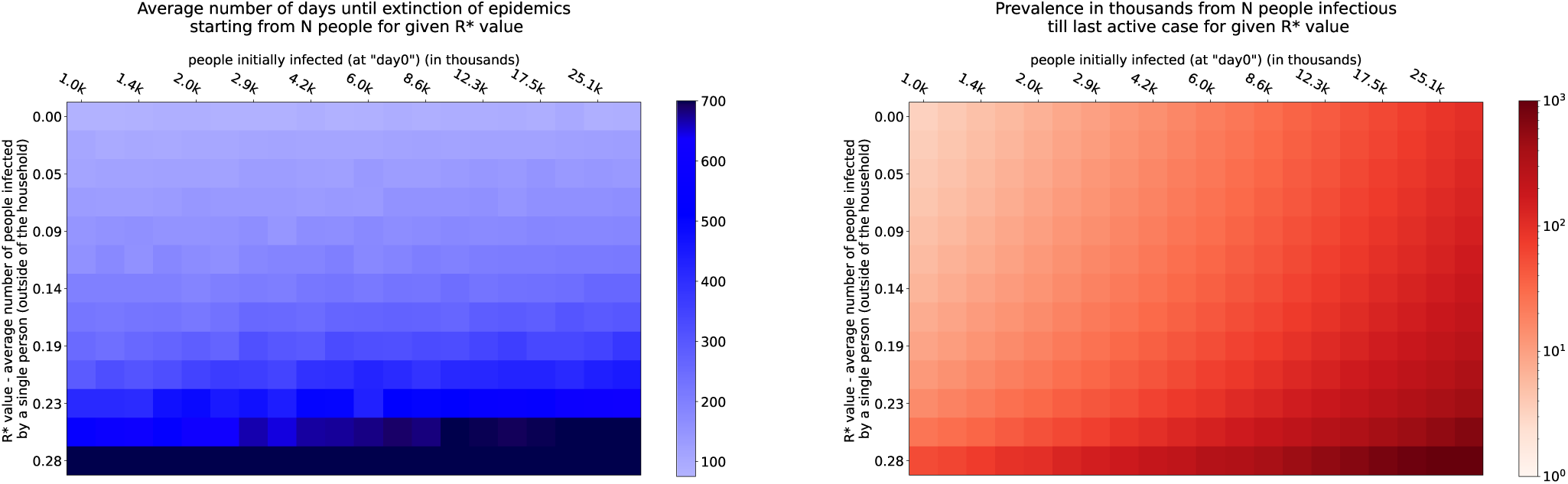
Figure 4a) Time till extinction in the subcritical regime in dependence of *R*^*^ and the initial number of infected individuals. Figure 4b) Prevalence in the subcritical regime in dependence of *R*^*^ and the initial number of infected individuals.

In Figure 5 we present results for additional scenarios which were only simulated for the city of Wrocław. Figure 5a shows for various parameters combinations of *R*^*^ and q - the probability for mild symptom patients to get tested - the prevalence after 200 days averaged over 10 independent simulations for each parameter pair. *R*^*^ refers here to the baseline scenario without quarantine, that is *R*^*^=2.34c. It should be noted that the true mean number of out-household secondary cases in the quarantine scenario is less than *R*^*^ and depends on q. The value q=0 corresponds to the base setting described above. Again, there is a rather narrow band of values for which mitigation is possible. In Figure 5b the blue colors indicate the average number of days after which the ICU threshold was surpassed. Parameter combinations where after 200 days less than 10 active cases were found are marked in white and correspond to subcritical progression. The yellow fields correspond to parameter combinations where neither of the first two criterion are fulfilled. For details see Appendix B. Therefore the parameter combinations for a sucessfull overcritical mitigation are limited within the yellow fields.

**Figure 5:**
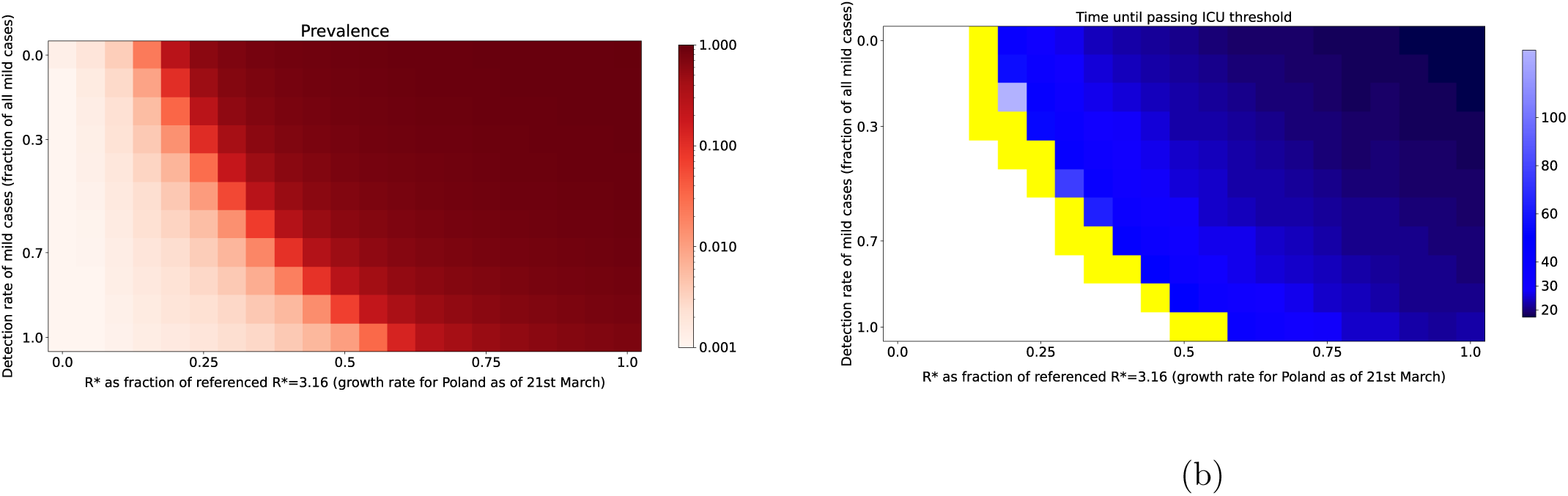
Epidemics outcome in Wrocław depending on a mixture of detection rate and social distancing measures (starting from 100 infections). Figure 5a displays the prevalence. In Figure 5b the blue colors indicate the average number of days after which the ICU threshold was surpassed. Parameter combinations where after 200 days less than 10 active cases were found are marked in white and correspond to subcritical progression. The yellow fields correspond to parameter combinations where neither of the first two criterion are fulfilled.

## 4. Conclusions

Semi-realistic microsimulations for Germany and Poland, on the basis of our model, give strong indications that there is only a narrow feasible interval of epidemiologically relevant parameters within which a successful mitigation is possible. Social distancing measures imposed by state authorities can hardly be fine-tuned enough to hit this critical interval precisely. Furthermore, herd immunity within these intervals is at best 15% percent of the population and would hence not provide sufficient protection for a second epidemic wave. The main reason for the narrowness of the mitigation interval as well as for the low critical value *R_min_* is the household structure. Infections within the households for patients with mild progression can hardly be avoided and therefore a small number of infection links between the households can already make the epidemic overcritical. In the subcritical domain we observe a strong dependence of time till extinction on the out-reproduction number *R*^*^. Reasonable extinction times are only achievable for very low values below 1/3 secondary infections outside of households.

It is of crucial importance to understand the source of the tightness of the phase transition as expressed by the narrowness of the gap between *R_m_in* and *R_m_ax.* Contrary to a microscopic model corresponding to a classical SEIR model based on ordinary differential equations, which is equivalent to an epidemic process on an Erdös-Renyi graph, the incorporation of households has a very strong impact on the location of the phase transition point and the steepness of the increase of the prevalence close to the critical value. Since the probability that a households of size *K* infects a household of size *L* follows a preferential attachment rule and hence is proportional to *K* · *L*, the variance, respectively the distribution of the household size matters rather than just the mean value, which is typical for non preferential couplings.

We conclude that instead, an extinction strategy implemented by quick, effective and drastic countermeasures similar to those put in action in China is ultimately required to reduce social contacts outside households and slow down the progression of the epidemic. If social distancing countermeasures are too weak there is a high risk of collapse of the public health system within a very short period of time. If contact reduction is not kept in force until disease extinction a second epidemic outbreak may result.^8^ Therefore, in order to control the epidemics it is nessesary to wait until it gets extinct. The application of an epidemic management plan based on a flawed strategy of herd immunity may easily lead to an uncontrollable epidemic. We also strongly advise combining social distancing and contact related countermeasures with an extensive testing strategy including individuals with characteristic symptoms but unknown contact history.

## Data Availability

The data we used is either open source or as the census data from the countries we used was provided from the state of Germany and Poland resp.

## 5. Acknowledgements

We would like to thank the University of Science and Technology Wrocław, the University of Wrocław and the Medical University of Wrocław for their support and assistance to the MOCOS group. We thank WCSS (Wrocaw Centre for Networking and Supercomputing) for giving us high priority access to computational resources. We also thank the City of Wrocław, Nokia Wrocław, EY GDS Poland and MicroscopeIT for their support. Special thanks goes to Dr. Jacek Pluta for his support and help for MOCOS on all administrative levels.

## Appendices

In Appendix A we give additional model specifications and additional timelines for various parameter settings. Detailed numerical values for the Figure 4 and Figure 5 are provided in Appendix B. In Appendix C we present and outline the equations used for the theoretic computation of the final prevalence.

### Appendix A. Model Specifications

#### Appendix A.1. Generation of the German synthetic households data

The German data was generated using the information gathered on household composition with the German Microcensus 2014^9^. For this purpose representative households were generated synthetically and resampled in order to fit the size of the population estimated from the same survey. To retain the regional heterogeneity of households across Germany, this was done for each of the federal states separately. Hence, also the composition of households for the Berlin data set is adapted to household structures in Berlin.

#### Appendix A.2. Generation of the Polish synthetic households data

The Polish data was generated using the data published by Statistics Poland (Gwny Urzd Statystyczny)^17,18^. The population was generated using data on the size and structure of the population by sex and age in all local and administrative units of the country as of 30 June 2019. The households for Poland were generated from the projection for year 2020 published in the Household projection for the years 2016–2050; each voivodeship was processed separately to preserve regional heterogeneity. The households for the city of Wrocław were generated based on the National Census of Population and Housing 2011^19^, Households and families in the Lower Silesian Voivodeship, and resampled in order to fit the size of the city population reported on 30 June 2019.

#### Appendix A.3. Fitting the *R*^*^ under the model to the observed infection cases

Under the assumption that the detection rate of infected persons is constant over time, the observed numbers of infected persons are valid for estimating the *R*^*^ in the population. To find the *R*^*^ under the present model that fits the observed evolution of cases in Germany and Poland and also for Wrocław and Berlin the following procedure was followed. First a very sparse grid of possible *R*^*^ was chosen to see the evolution of the stochastic microsimulation over time. As the observed data from 5th March till 20th March were available, the increase from a starting number of infected persons to the final number, as of 20th March, should have occurred within these 16 days. To show the narrowness of the possible parameters an example for this procedure will be made for the federal state of Berlin. In Figure A.6 the box-plots of the duration are plotted for obtaining an increase of 13 (5th March, Berlin) to 848 (20th March, Berlin) in 16 days. Values above the horizontal line at 16 indicate that *R*^*^ is too low, as it takes too long to take the evolution, and values below 16 state that *R*^*^ is too high, as it evolves faster than the observed values. However, as we are dealing with a stochastic microsimulation it is clear that the outcome after a given time is not deterministic, but may vary quite a lot. Therefore, the dots indicate the average duration to perform the above said evolution. From this Figure A.6 the suspected *R*^*^ could be seen to be somewhere in the neighbourhood of 3.883. To validate this, we then look at the different runs and compare it with the observed values in a semilog plot as is usual for exponential growth functions. This plot is given for Berlin in Figure A.7.

**Figure A.6:**
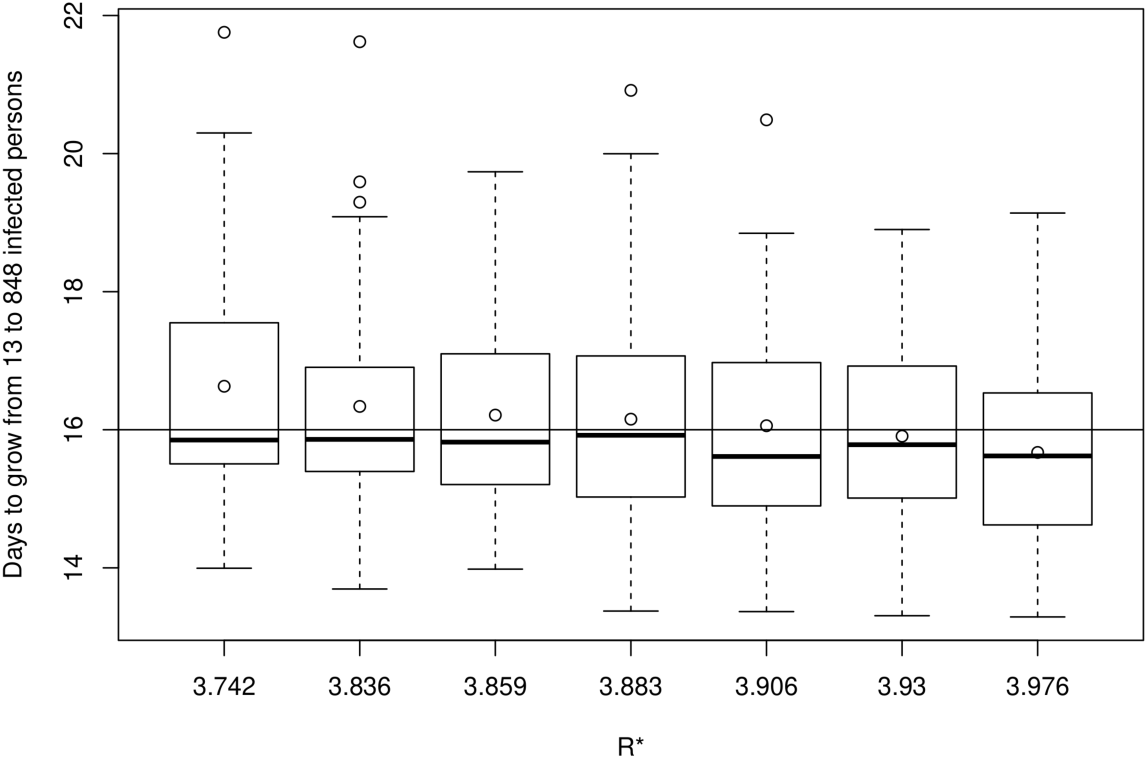
Boxplot of time needed in the simulation to reach 848 infected people starting form 13 infected people in Berlin given different *R*^*^ parameters.

**Figure A.7:**
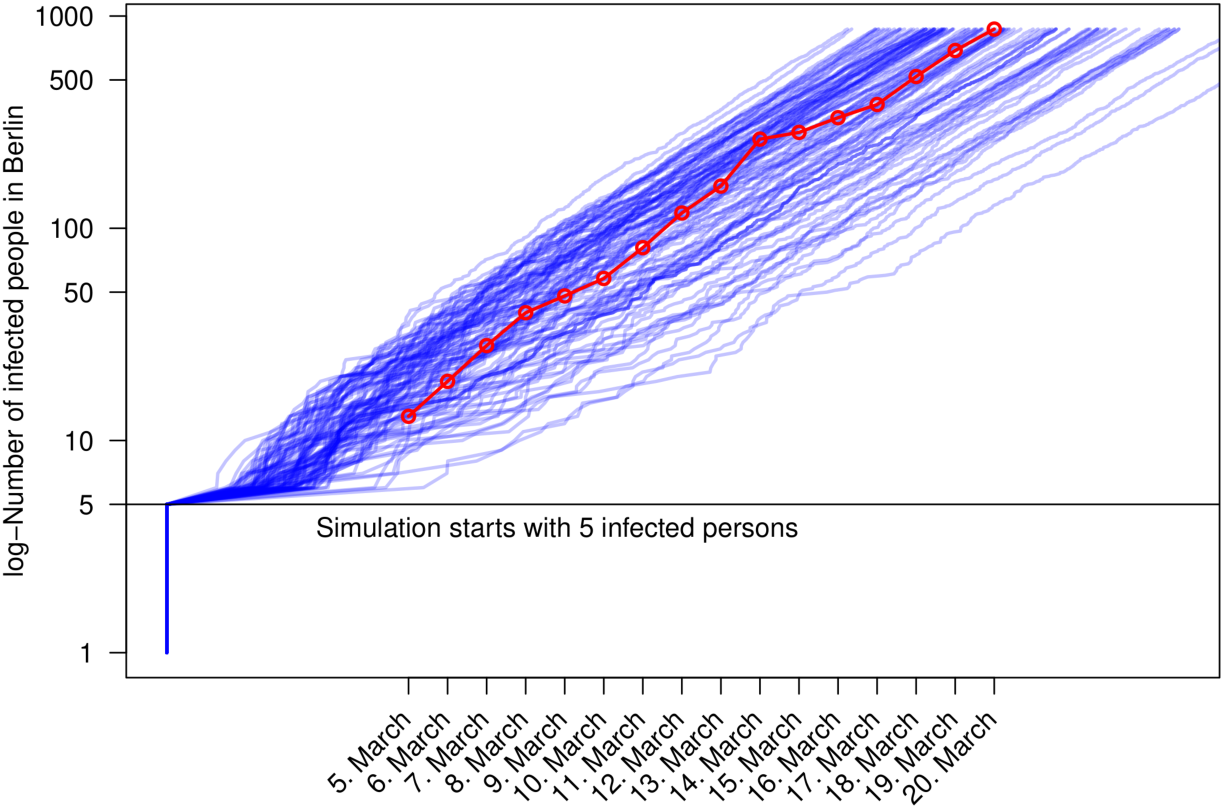
Development of the epidemic in Berlin given the fitted *R*^*^ of 3.883. Blue lines are developments under different random runs and red dots show the observed number of infected in the timespan of 5th March to 20th March.

Each blue line represents one simulation run. The simulation started with 5 infected persons. As can be seen, as usual in stochastic microsimulations, it takes some time till the dynamic of the simulation stabilizes, which is definitely already the case when 13 infected persons are reached in the average of runs. This point is indicated as the 5th March where this number of infected individuals was observed in Berlin. As can be seen from the graph, the red line indicating the observed values lies in the center of the simulation path. So we are confident that this *R*^*^=3.883 is a good approximation to the actual *R*^*^ in Berlin. Similarly, the *R*^*^ are gathered for Poland *R*^*^=3.159 (5th of March until 20th of March) and Germany *R*^*^=3.041. Note that the epidemic in Wrocław is still in such an early stage that the case numbers do not yet allow a reliable estimation. We thus assume for now that the *R*^*^ for Wrocław coincides with the estimated *R*^*^ for Poland.

#### Appendix A.4. Searching for the *R_min_* and *R_max_*

In contrast to *R*^*^ there is no data we can align the simulation to for obtaining the *R_min_* and *R_max_* values. We defined *R_max_* as the smallest *R*^*^ value for which 10 sample paths surpassed the ICU threshold within D days. For cities D was chosen to be 200 days and for countries 700 days. The critical value *R_min_* was defined as the largest *R*^*^ < *R_max_* for which the daily incidence at day 200 was below 50% of the initial number *N_o_* of infected. That is, it represents the *R*^*^ that will most probably not threaten the health-care system. To find these values we started fitting a sparse grid of plausible *R*^*^. By bisection we gradually made the grid finer to approach both *R_min_* and *R_max_* as described above.

**Figure A.8:**
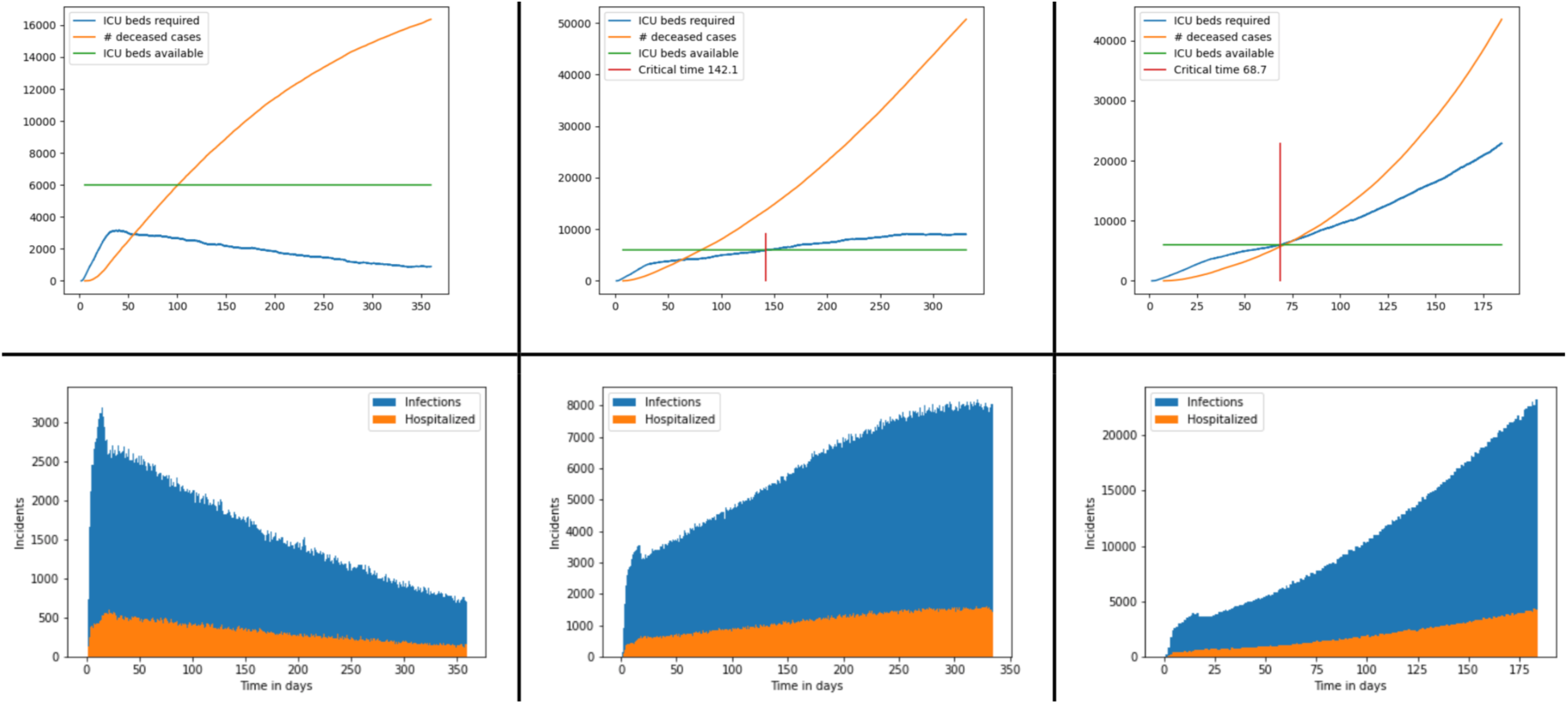
The progress of the epidemic for *R_min_* (left), *R_max_* (right) and one value in between for Germany.

**Figure A.9:**
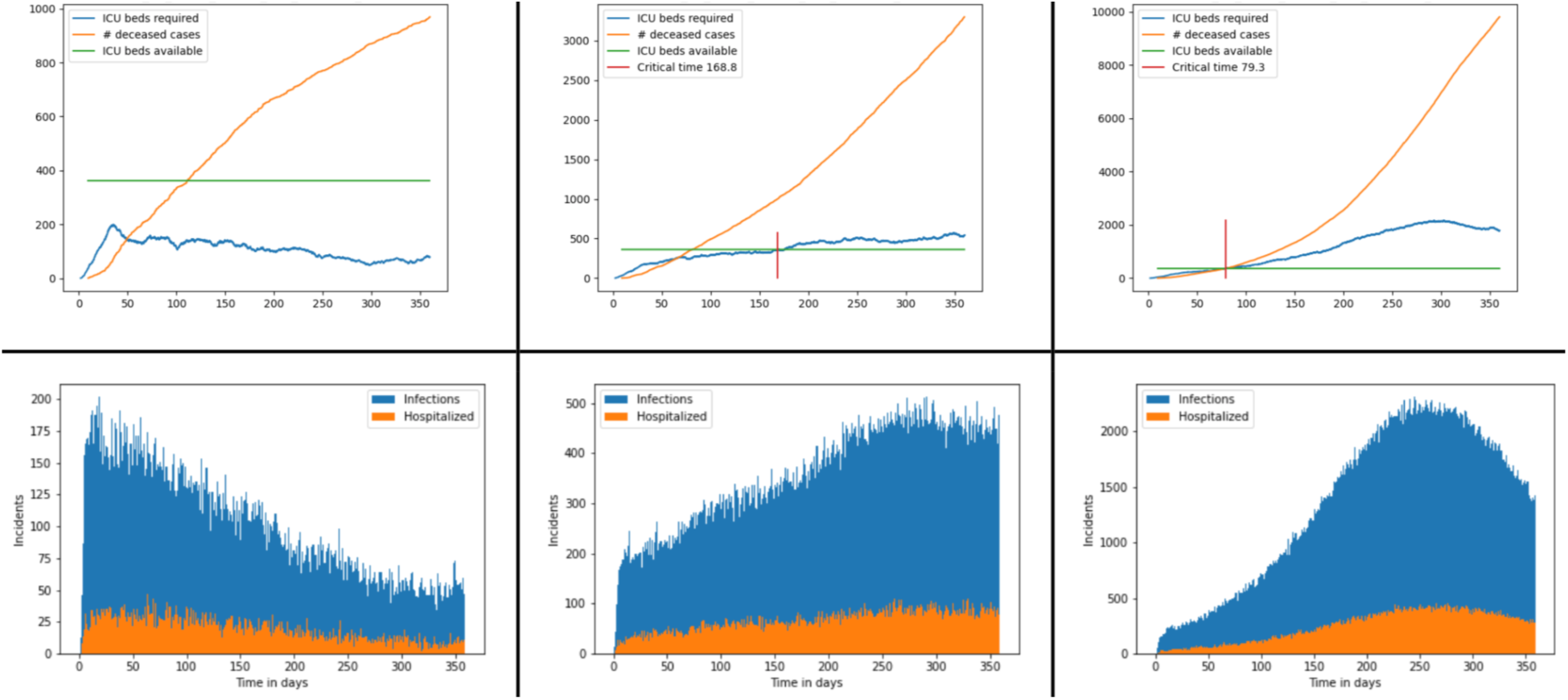
The progress of the epidemic for *R_min_* (left), *R_max_* (right) and one value in between for Berlin.

### Appendix B. Detailed Values for Figure 4 and Figure 5

Figure B.10 gives the numerical values for the time till extinction in the subcriticial regime.

**Figure B.10:**
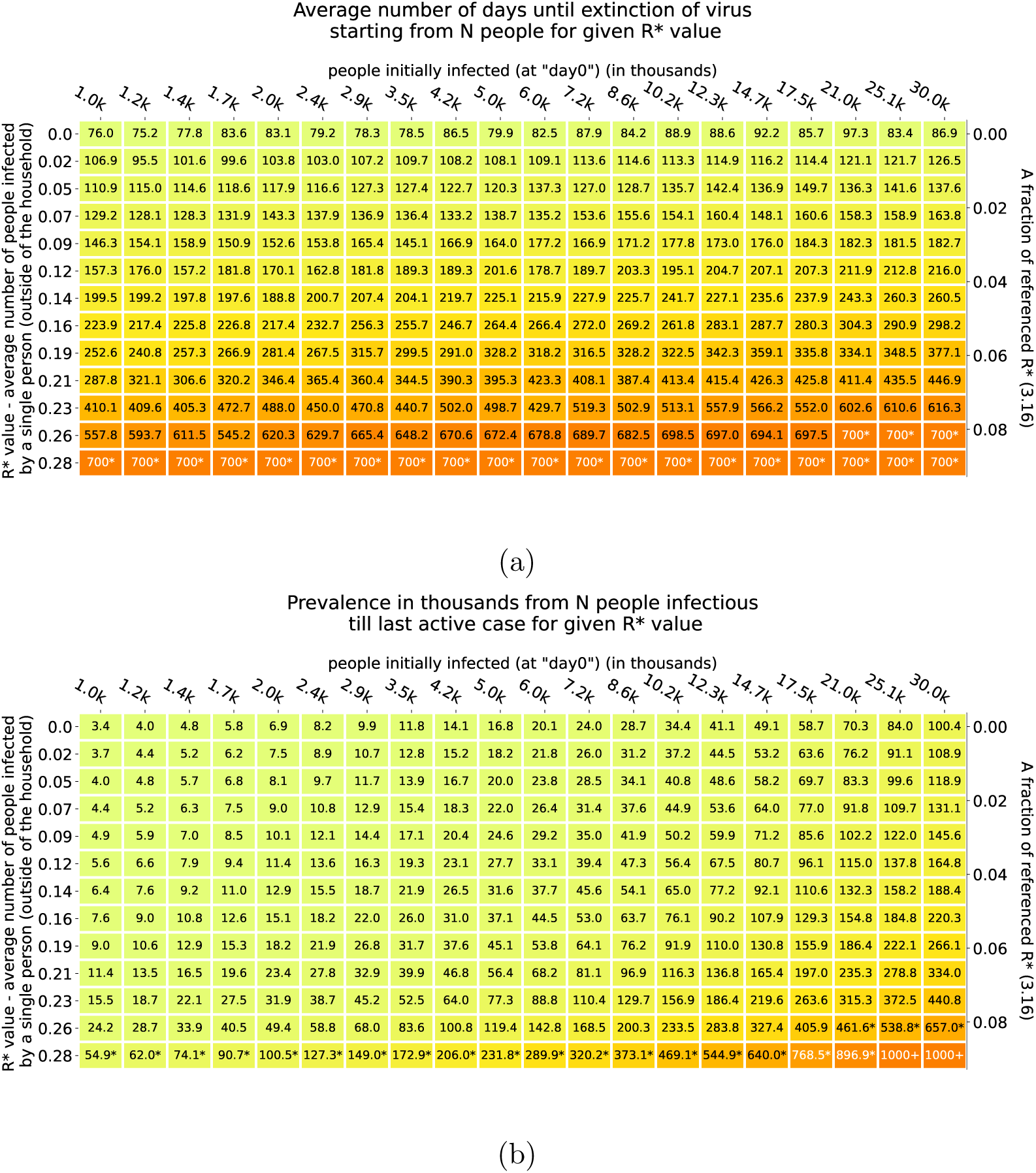
a) shows time till extinction of the epidemics for Poland for selected *R*^*^ values and different initial numbers of those infected. Simulations were stopped at 700 days if time till extinction was longer or at 1 million infected people if prevalence was larger. Figure 4b) shows the corresponding prevalences at the end of the simulation in thousands. Reported times and prevalences are averages over 10 simulations for each parameter combination. For each simulation the initial infected individuals were uniformly sampled from the population. Numbers in the bottom denoted with * are due to the fact that epidemics were not stopped in 700 days. Additionally, two last numbers were labelled as 1000+ as the second stopping criterion (prevalence > 1 million) was triggered.

In Figure B.11 we present numerical results for Figure 5. Parameter combinations where after 200 days less than 10 active cases were found are marked in green and correspond to subcritical progression (for fields with percentage numbers not all simulations ended with less than 10 active cases; the percent number refers to the fraction of simulations for which less than 10 active cases at 200 days were observed). In Figure B.12 we display by red fields with numbers those parameter pairs for which the ICU demand surpasses the threshold of ICUs (for fields with percentage numbers not all simulations ended with less than 10 active cases; the percent number refers to the fraction of simulations for which less than 10 active cases at 200 days were observed). The average number of days after which this happens is presented in the same figure. A red field with no number in can be considered as a pair of parameters for which successful mitigation takes place. The white fields correspond to overcritical parameter combinations for which the whole epidemic has already finished and less than 10 active cases were found 200 days after the outbreak.

**Figure B.11:**
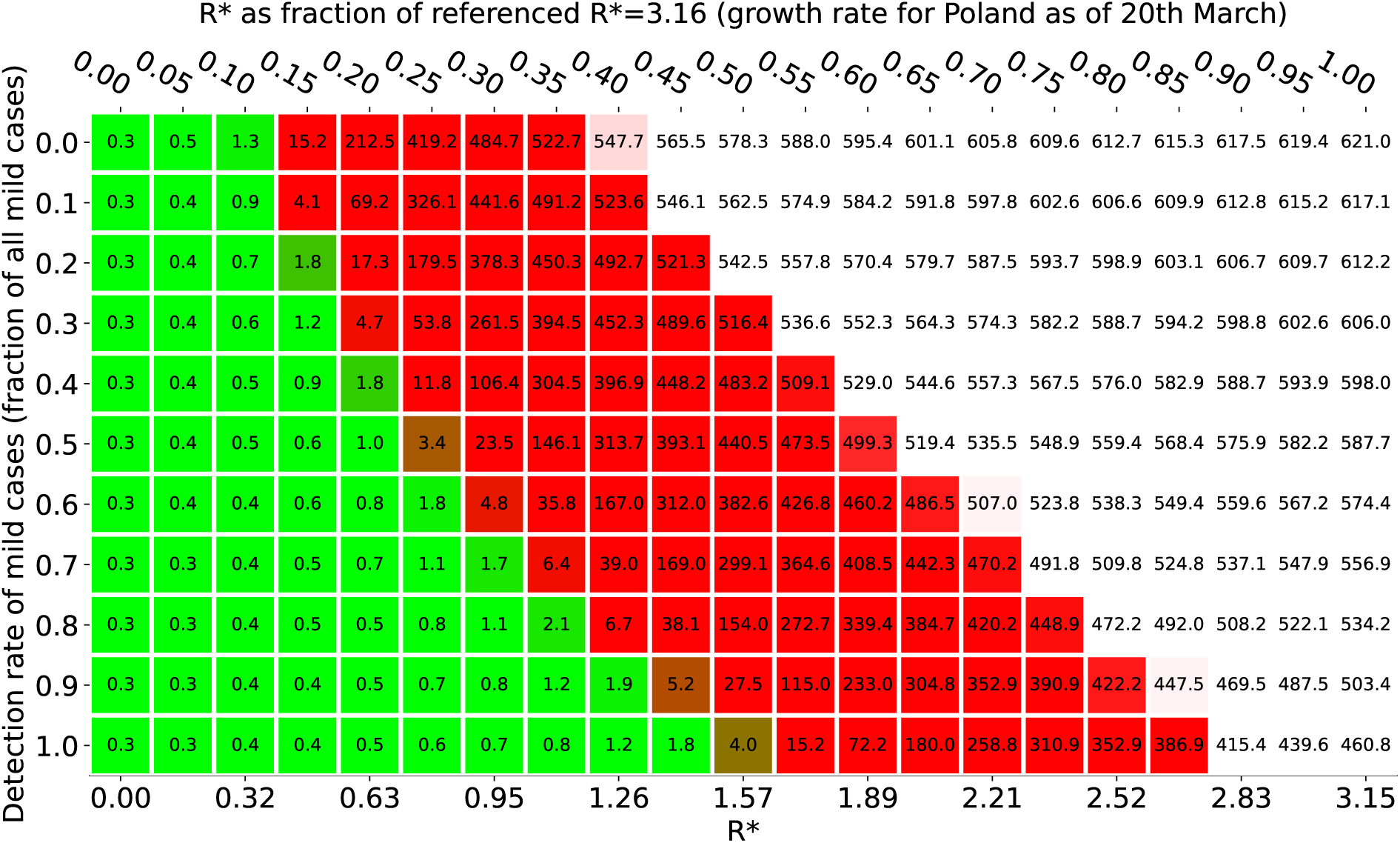
Epidemics outcome in Wrocław depending on a mixture of detection rate and social distancing measures (Starting from 100 infections). Green color indicates a subcritical regime (less than 10 cases after 200 days); Red color indicates a critical regime; White color indicates a ‘supercritical’ regime (full blown epidemics ends within 200 days). Numbers in Figure a) denote epidemics prevalence after 200 days; numbers are given in thousands.

**Figure B.12:**
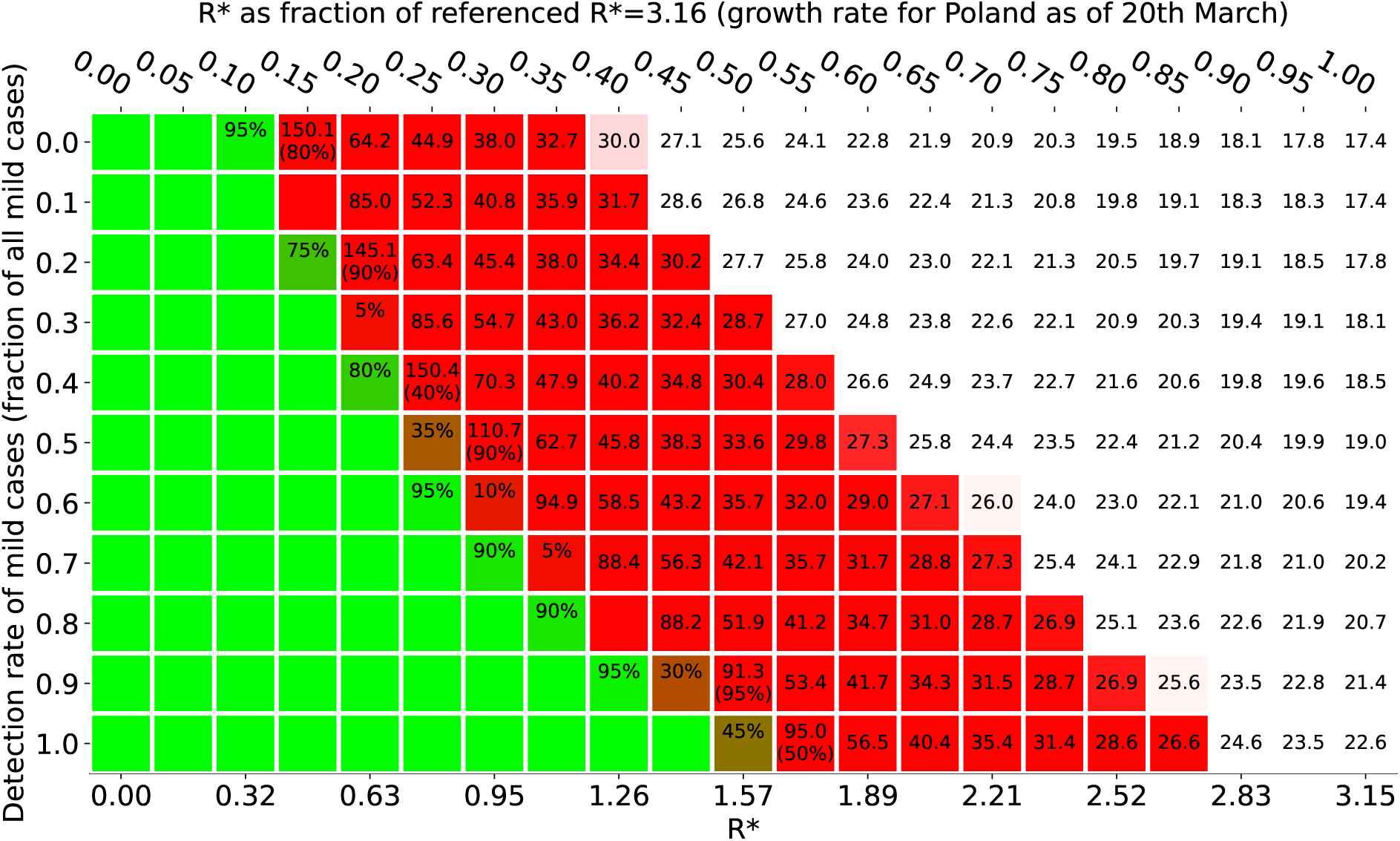
Epidemics outcome in Wrocław depending on a mixture of detection rate and social distancing measures (Starting from 100 infections). Numbers in red fields denote the mean number of days it takes until the ICU capacity is exceeded. Percentage number in green fields give the fraction of simulations for which less than 10 active cases after 200 days were observed and percentages in red fields give the fraction of simulations which exceeded the ICU capacity threshold. All averages were taken over 10 simulations.

### Appendix C. Mathematical Supplement

The theoretical curve in Figure 2 was computed on the basis of results from the theory of heterogenous random graphs^20^ and multitype branching processes. We adapted known formulas for the size of the giant component in sparse heterogenous random graphs. To do so we had to take into account the clique structure within households and the specific disease progressions within patients given by our model setup. We first evaluated the size the giant component on a random graph representing the infectious connections between household. The final prevalence of the infection in the population can the be obtained by a proper weighted scaling. In detail we used the following formulas. We denote by *h*(*k*) the proportion of households of size *k* in our sample population, hence 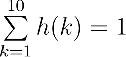. Let further 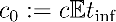 be the average number of people an individual infects outside the household where 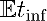 is the expected time being infectious outside the household. Let further 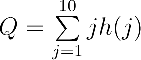 be the average household size. The constant *c* is the intrinsic parameter described in the model description of the main text. Then fraction *ρ(k)* of infected households in the giant component is given by the largest solution of

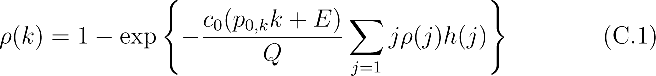

where *p*_0_,*_k_* is the probability that a household member in a *k*− size household will not infect any other household member and *c*_0_*E* is the expected number of secondary cases generated by an infected individual outside it’s household conditioned that the individual will have a disease progression which makes a hospital stay necessary.

Here *ρ*(*k*) can also be interpreted as the survival probability of an associated branching process describing the initial spread of the epidemics starting with one infected household of size *k*. The quantities *p*_0_,*_k_* and *E* in the exponent take into account the combined effect of reduced household infections due to patients which get early hospitalized and the reduced number of secondary infections generated by such patients. The numerical values of *p*_0_,*_k_* and *E* can be computed from the underlying distributions for disease progression within patients.

The relative prevalence of the epidemics in the sample population is then given by

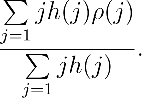

Note that the formulas estimate only asymptotic relative size of the giant component and therefore cannot be used to estimate prevalences for subcritical epidemic.

Details, further refinements and proofs will appear in a forthcoming paper.

**Figure C.13:**
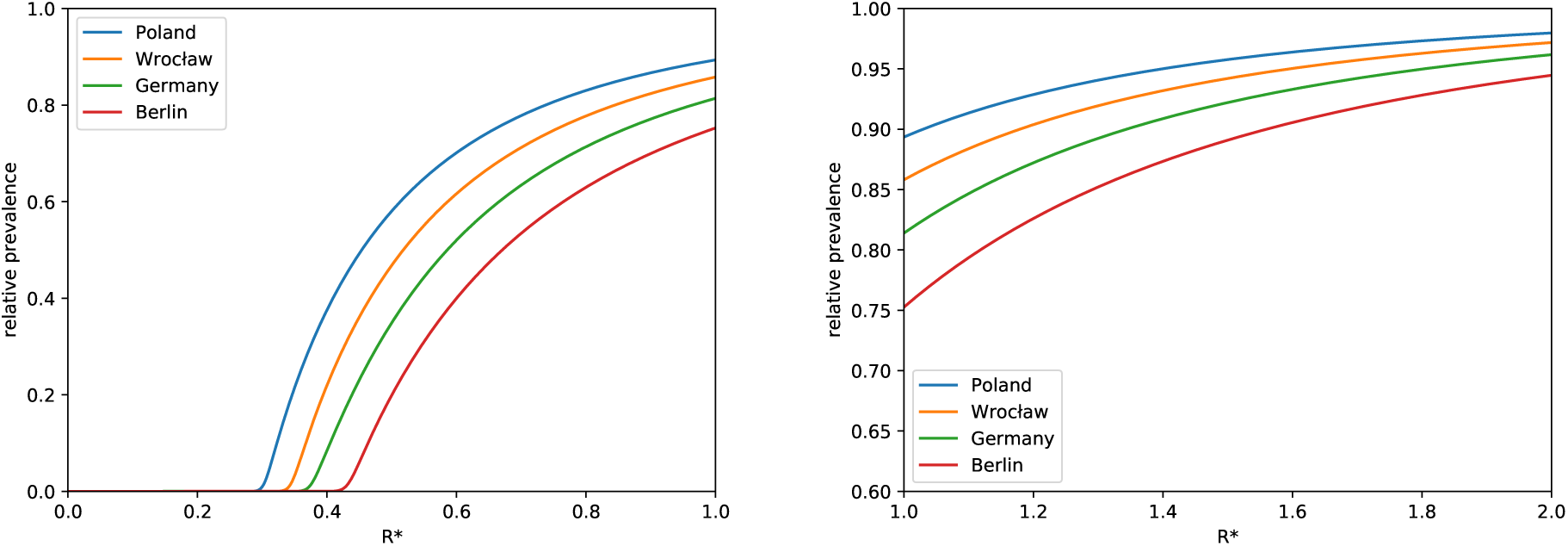
Theoretical predictions for the relative end-prevalence of the epidemic as a function of *R*^*^ for *R*^*^ ≤ 2. The differences between the two countries and cities are due to the different household size distribution.

The critical value of *c* for which the epidemics becomes overcritical (respectively the giant component emerges) is characterised by the value where spectral norm of the associated transfer operator, for underlying branching process equals one. The values for which the norm is larger than one hence correspond to the overcritical region.

**Figure C.14:**
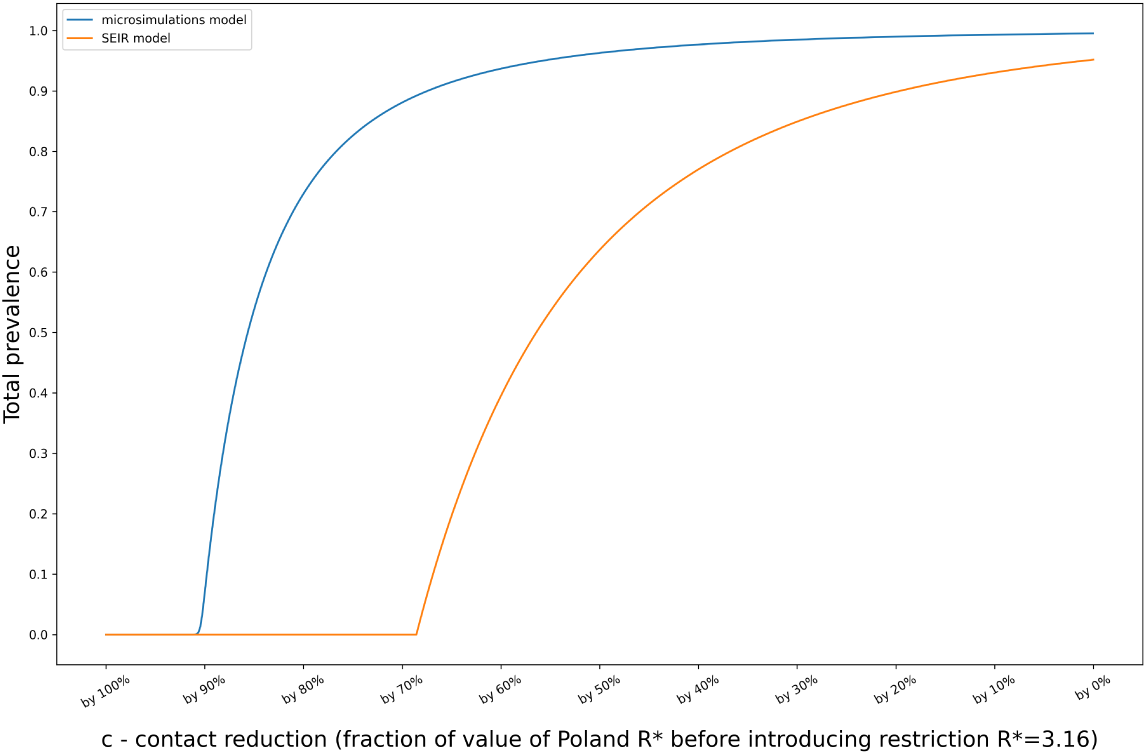
Total prevalence as a function of the contact reduction. A comparion between an SEIR model and the model used throughout this paper.

